# Development and Evaluation of Deep Learning Models for Cardiotocography Interpretation

**DOI:** 10.1101/2024.03.05.24303805

**Authors:** Nicole Chiou, Nichole Young-Lin, Christopher Kelly, Julie Cattiau, Tiya Tiyasirichokchai, Abdoulaye Diack, Sanmi Koyejo, Katherine Heller, Mercy Asiedu

## Abstract

The inherent variability in the visual interpretation of cardiotocograms (CTGs) by obstetric clinical experts, both intra- and inter-observer, presents a substantial challenge in obstetric care. In response, we investigate automated CTG interpretation as a potential solution to enhance the early detection of fetal hypoxia during labor, which has the potential to reduce unnecessary operative interventions and improve overall maternal and neonatal care. This study employs deep learning techniques to reduce the subjectivity associated with visual CTG interpretation. Our results demonstrate that using objective umbilical cord blood pH outcome measurements, rather than clinician-defined Apgar scores, yields more consistent and robust model performance. Additionally, through a series of ablation studies, we explore the impact of temporal distribution shifts on the performance of these deep learning models. We examine tradeoffs between performance and fairness, specifically evaluating performance across demographic and clinical subgroups. Finally, we discuss the practical implications of our findings for the real-world deployment of such systems, emphasizing their potential utility in medical settings with limited resources.

## 1 Introduction

Intrapartum cardiotocography (CTG) is a screening technique that is widely used to monitor fetal well-being by recording the fetal heart rate (FHR) along with the maternal uterine contractions (UC) during labor. Although CTG is routinely used in medical practice, the use of continuous intrapartum fetal monitoring is associated with a high false-positive rate. This has led to an increase in Cesarean section and operative vaginal delivery rates with limited improvements in neonatal outcomes [1] due to the subjectivity of current methods [2, 3] and intra-observer variability [4]. These issues are exacerbated in low-resource facilities where access to skilled interpreters is limited [5, 6]. Figure 1 highlights the aforementioned challenges with visual CTG interpretation and the relevant clinical use case.

**Figure 1.**
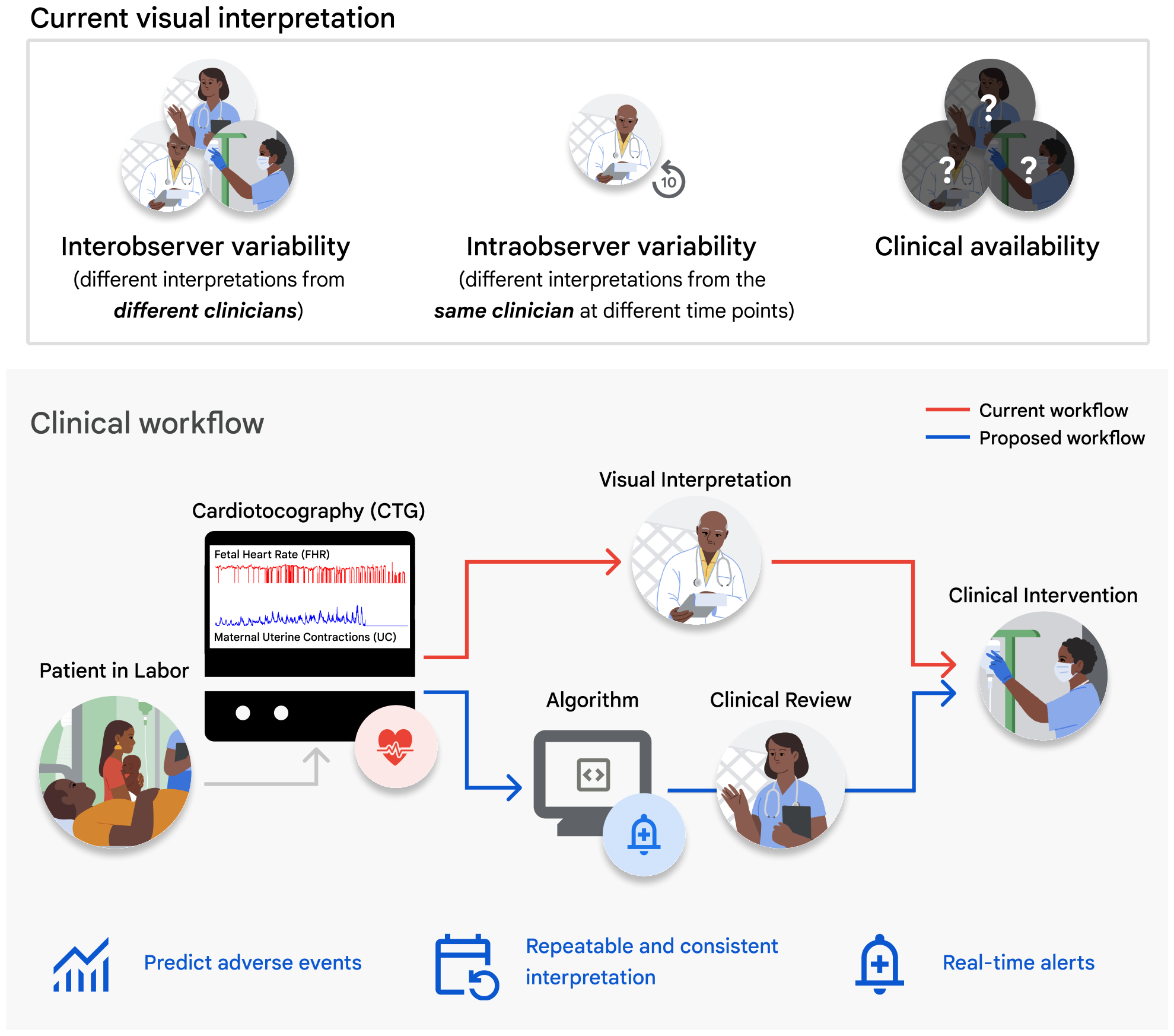
*Top*: Challenges of current visual CTG interpretation. *Bottom*: Proposed clinical use case of deep learning algorithms for CTG interpretation and assistive clinical decision-making.

Machine learning algorithms to classify abnormal CTGs from tabulated rules-based extraction of diagnostic features have shown promise for improving clinical decision support [7–12]. However, they reduce rich CTG signal information to a few numbers which ignore important temporal and contextual cues such as the relative timing of delivery, maternal risk factors, etc. [13–16].

Current deep learning methods for CTG interpretation, which use the physiological time series data as input, rely on proxy labels for fetal well-being recorded immediately after delivery: the umbilical artery blood pH and the 1-minute Apgar score [17–23]. Umbilical cord blood pH at the time of birth, often used in high-resource medical facilities, is presently the only objective quantification for the potential occurrence of fetal hypoxia during labor. In contrast, the Apgar score at one minute from delivery is a subjective score from 0 to 10 assigned by a clinician and reflects the general health of the newborn. Apgar scores are the primary delivery outcome descriptor in low-resource settings due to their simplicity, cost-effectiveness to adopt, and the potential financial burden of umbilical blood analysis [24, 25].

Additionally, while CTG is widely recorded continuously in high-resource facilities, various system and implementation challenges faced by low- and middle-income countries (LMICs) constrain recording to intermittent periods, which may often exclude the signal interval immediately preceding delivery [26–28]. Thus, to enable applications in low-resource clinical use cases, machine learning-based solutions must aim to accurately detect fetal compromise at arbitrary time points before delivery to enable timely obstetric intervention [26, 28, 29].

### Contributions

- We highlight the feasibility of using deep learning methods to reduce the subjectivity of predicting fetal hypoxia from visual CTG interpretation. We conduct ablation studies to analyze the effect of (a) the choice of objective (pH) vs subjective (Apgar) ground truth labels, (b) the signal time interval used for training, and (c) the evaluation of simulated low-resource environment signals on predictive performance.
- We propose data augmentation and statistical evaluation methods to overcome challenges with this limited dataset.
- Finally, we discuss the implications of training deep learning models for deployment in low-resource settings from a global health perspective.

## 2 Methods

### 2.1 Dataset description

The CTU-UHB Intrapartum Cardiotocography Database is an open-source selective collection of 552 CTGs consisting of *∼*50,000 minutes of recordings at term (*≥* 37 weeks gestation) [30, 31]. Each CTG records the fetal heart rate (FHR) and corresponding uterine contractions (UC) for up to 90 minutes before delivery. The data are associated with fetal outcomes, along with fetal and maternal metadata, summarized in Table 1. A correlation matrix of the associated metadata is included in Figure 2A. This dataset was retrospectively accessed and did not require IRB approval. We defined three outcome label categories:

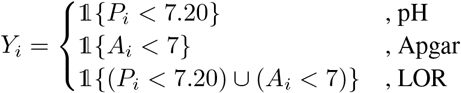

where 1{*·*} is the binary indicator function, *P*_*i*_ is the umbilical arterial cord blood pH, *A*_*i*_ is the 1-minute Apgar score, and *Y*_*i*_ is the assigned ground truth label (abnormal = 1, normal = 0) for the *i*th CTG recording. The pH classification task yielded 375 normal and 177 abnormal cases, with a cut-off threshold of 7.20. This threshold was chosen for relevance to clinical cut-offs, appropriate class balance, and to enable comparison to prior work on a similar task [19]. The Apgar classification task yielded 484 normal and 68 abnormal cases. The LOR classification task, defined as the logical inclusive “OR” of the abnormal pH and Apgar criteria yielded 354 normal and 198 abnormal cases [19]. Examples of normal and abnormal CTGs are shown in Figure 2C. Figure 2D shows the relationship between pH and Apgar scores for all patient recordings.

**Table 1:** Patient metadata, CTG recording, and labor outcome descriptors for the CTU-UHB dataset. The mean, median, and/or range are shown for continuous variables. The approximate total duration of the CTG signal, in minutes, and the number of unique recordings (count) are shown for the remaining binary variables.

|  | Mean (Median) [Range] |
| --- | --- |
| Maternal age (years) | 29.8 [18, 46] |
| Parity | 0.43 (0) [0, 7] |
| Gravidity | 1.43 (1) [1, 11] |
| Gestational age (weeks) | 40 [37, 43] |
|  | Approximate minutes of CTG (Count) |
| Diabetes | 1,100 (37) |
| Hypertension | 1,320 (44) |
| Pre-eclampsia | 510 (17) |
| Liq. praecox | 4,380 (146) |
| Pyrexia | 180 (6) |
| Meconium | 1,920 (64) |
| Induced | 6,510 (217) |
| Vaginal delivery | 15,180 (506) |
| C-section | 1,380 (46) |
| External FHR monitoring | 12,390 (413) |
| Internal FHR monitoring | 3,060 (102) |
| External and internal FHR monitoring | 1,050 (35) |
| Umbilical artery pH < 7.20 | 5,310 (177) |
| 1-minute Apgar < 7 | 2,040 (68) |
| LOR (pH < 7.20) ∪ (Apgar < 7) | 5,940 (198) |

**Figure 2.**
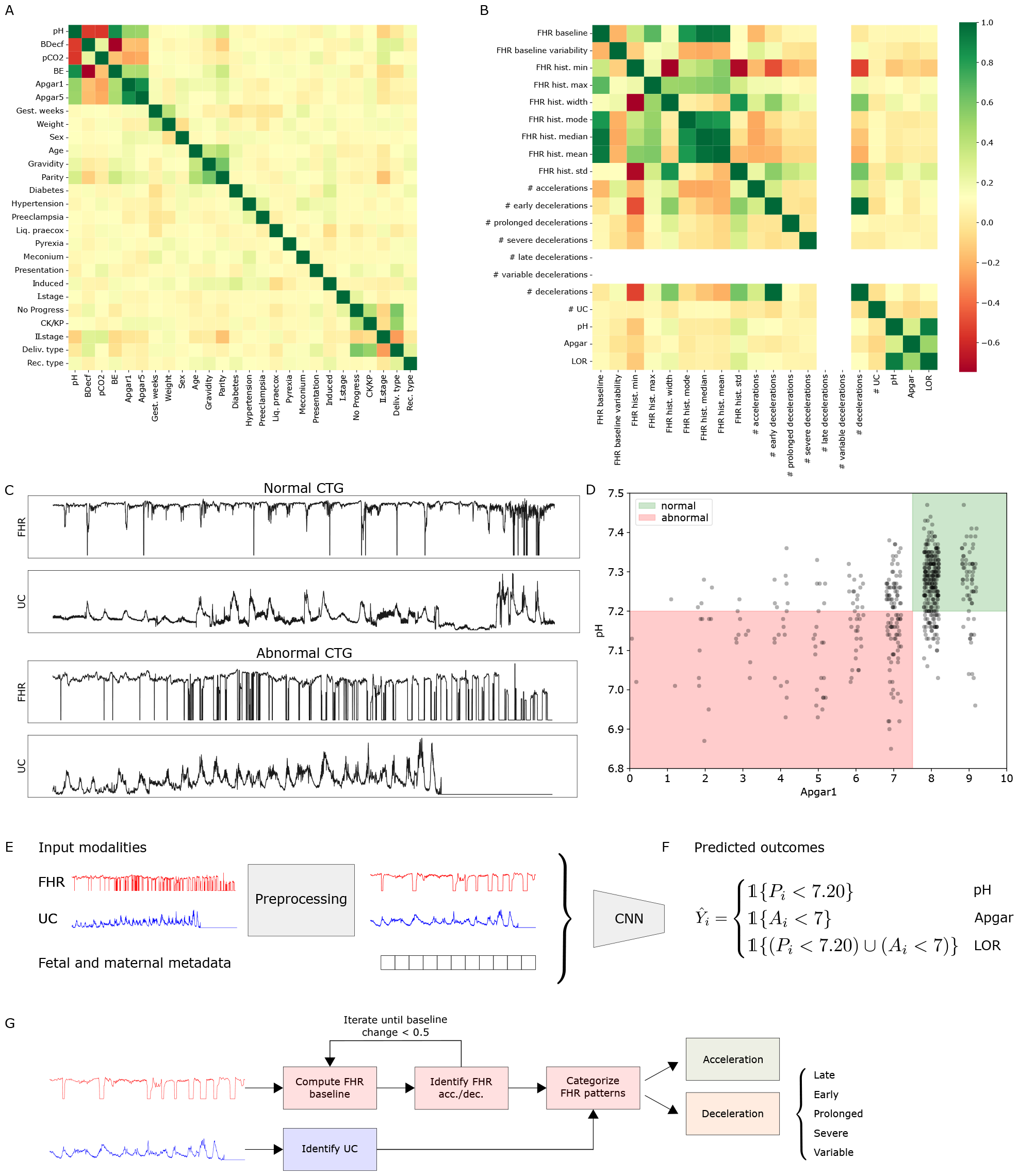
(A) Correlation between patient metadata attributes. (B) Correlation between extracted features and predicted target variables. (C) *Top*: Normal CTG recording with pH *≥* 7.20 and Apgar at 1-minute *≥* 7. *Bottom*: Abnormal CTG recording with pH *<* 7.20 and Apgar at 1-minute *<* 7. (D) pH and Apgar scores for all patient CTG recordings. The green region denotes the intersection of normal recordings and the red region denotes the intersection of abnormal recordings. This figure demonstrates that there is not necessarily a direct link between pH values and Apgar scores. (E) Possible input modalities for the deep learning model, which outputs (F) a predicted outcome depending on the classification task. (G) Rules-based feature extraction pipeline. model for 300 epochs [34]. We perform channel-specific maximum absolute value scaling on time series and metadata normalization on tabular features before input into the neural network.

### 2.2 Preprocessing for the neural network methods

The preprocessing pipeline for signals input into the neural network models included removing repeated zero signals at the beginning and end of the recorded signal, data quality assessment, imputation of missing values, signal smoothing, data augmentation, cropping to 30-minute signal length, and downsampling. This resulted in 4,315,200 minutes (*n* = 496), 148,800 minutes (*n* = 496), and 1,680 minutes (*n* = 56) of signal for pre-training, training, and testing, respectively. Details of the preprocessing steps can be found in Appendix A.

### 2.3 Alternative time interval preprocessing

#### Training on different time points but evaluating on last 30 minutes of the held-out set

To train models on cropped recordings from different time intervals we used the following: (1) cropped signal with a sliding window of a 30-minute interval and 1-minute step size that excluded the last 30 minutes of signal and (2) randomly cropped 30-minute windows sampled over the entire recording. (3) We also investigated pre-training on signals from (1) and then fine-tuning model parameters on the last 30 minutes of the signal in the training set. We partitioned the CTG recordings into identical disjoint training and testing sets for both the pre-training and fine-tuning phases to ensure that none of the patient recordings in the held-out test set were previously seen by the fine-tuned model during pre-training.

#### Training on the last 30 minutes but evaluating different time points of the held-out set

We evaluated the baseline model trained on the last 30 minutes of signal on the following varied 30-minute segments in the held-out dataset: (1) the signal at the 30-60 minute mark before delivery and (2) randomly cropped 30-minute signals sampled over the entire recording. These time intervals simulate the type of observed signal common in resource-constrained settings. These samples had no additive multi-scale noise and were generated in a fully deterministic fashion.

### 2.4 Data splitting

Due to the imbalance of classes and small patient sample size in the CTU-UHB dataset, we performed stratified data splitting to ensure similar distributions over the predicted target variable across training, validation, and test splits. Since the augmented dataset had a one-to-many correspondence between record identifiers and augmented samples, we first assigned 10% of the record identifiers to a held-out test set before assigning the remaining 90% to 10-fold cross-validation (cv) splits by record identifiers, ensuring a representative distribution of labels in each split.

### 2.5 Neural network model architecture and training

We utilized the CTG-net neural network model proposed by Ogasawara et al. [19] for our base model. The CTG-net architecture takes signals of 1800 time points long (30 minutes downsampled at 1 Hz) as input. The input FHR and UC signals are convolved with 30-second temporal filters before a depthwise convolution is conducted to learn the relationship between FHR and UC. A final separable convolution is applied before all features are flattened and passed to fully-connected hidden layers for classification. The output of the final layer is passed through a sigmoid activation layer to yield an abnormality score. A high-level depiction of the training pipeline is shown in Figures 2E-F. Further details regarding the architecture can be found in Ogasawara et al. [19].

We also ran the following experiments: (1) training with FHR or UC as input using a 1D CNN model variation and (2) adding metadata features as a vector to the input. We performed hyperparameter tuning through an architecture search. Model hyperparameters were optimized separately for two-channel (FHR and UC) versus 1-channel (FHR) input models. Out of 500 random hyperparameter configurations, the best hyperparameters were selected based on the highest validation AUROC averaged over 10 cross-validation folds. See Appendix B for more details.

All neural networks were trained on an NVIDIA V100 GPU in TensorFlow using the Keras API [32, 33]. We used Adam as the optimizer, initialized model weights and optimizer state with a fixed random seed, and trained each

### 2.6 Rules-based classification with XGBoost

To compare the performance of the neural network model to conventional machine learning algorithms, we implemented a feature extraction pipeline to extract FHR and UC features from the time series recording according to the current maternal and fetal medicine practices of the International Federation of Gynecology and Obstetrics (FIGO) [35]. The following features were extracted: uterine contractions, FHR baseline, baseline variability, accelerations, decelerations, variable decelerations, severe decelerations, late decelerations, prolonged decelerations, and the width, minimum, maximum, median, mean, mode, and standard deviation values of the FHR histogram. Figure 2B shows the correlation matrix for all extracted features and predicted target variables. Figure 2G depicts the feature extraction pipeline. Further method details can be found in Appendix A.

We trained an XGBoost classifier, typically used in CTG applications [8], on the extracted FHR and UC features for the pH and Apgar classification tasks using the same data splits used to train the neural network models. We used 1000 estimators with a maximum depth of 2, a class-corrected weighted loss, 30 early stopping rounds, and AUROC as the evaluation metric.

### 2.7 Evaluation

A summary of all conducted experiments can be found in Table 2. The performance of the neural network and XGBoost models was evaluated using the area under the receiver operating characteristic curve (AUROC). We also reported the sensitivity at a fixed specificity threshold of 90% for comparison with clinical performance.

**Table 2:** Summary of experiments conducted for the pH and Apgar prediction tasks. ^*∗*^ denotes the inclusion of additional experiments for the LOR prediction task, ^*†*^ indicates the execution of a subgroup analysis, and ^*‡*^ indicates completion of a single attribute metadata sweep.

| Inputs | Train time interval | Test time interval |
| --- | --- | --- |
| Feature-based | Last 30 minutes | Last 30 minutes |
| UC | Last 30 minutes | Last 30 minutes |
| FHR | Last 30 minutes | Last 30 minutes<br>Last 30-60 minutes<br>Random 30 minutes |
|  | Pre-train<br>Random 30 minutes<br>Pre-train + fine-tune | Last 30 minutes |
| FHR + metadata | Last 30 minutes | Last 30 minutes |
| FHR + UC | Last 30 minutes | Last 30 minutes*<sup>†</sup><br>Last 30-60 minutes<br>Random 30 minutes |
|  | Pre-train<br>Random 30 minutes | Last 30 minutes<br>Last 30 minutes |
|  | Pre-train + fine-tune | Last 30 minutes<br>Last 30-60 minutes<br>Random 30 minutes |
| FHR + UC + metadata | Last 30 minutes | Last 30 minutes<sup>†‡</sup><br>Last 30-60 minutes<br>Random 30 minutes |

A two-tailed Welch’s *t*-test was used to compare the average AUROC computed over bootstrapped samples for the various approaches. In cases where we compared performance on a fixed test set, a paired *t*-test was used. We also evaluated performance disparities across subgroups for both pH and Apgar prediction tasks. Subgroup variables included demographic and clinical attributes (gestational age of the fetus, maternal age, maternal risk factors, parity) as well as signal quality descriptors for each of the FHR and UC channels. A comprehensive description of the cut-offs and formulas used to define binary subgroup variables is found in Appendix A.

## 3 Results

### 3.1 Evaluating model performance by prediction task, input signal type, and metadata annotation

#### Performance by prediction outcome (pH vs. Apgar and LOR ground truth labels)

Our baseline method for predicting LOR takes both FHR and UC signals and achieves comparable AUROC (0.68 *±* 0.07) as prior work (0.68 *±* 0.03) [19]. We achieved a lower (0.27 *±* 0.18) sensitivity at 90% specificity than clinician performance (0.45, 95% CI: 0.23-0.68) reported in an observational study with 500 patients, however, this was not significant [36]. Baseline Apgar prediction was slightly higher (0.69 *±* 0.12) and baseline pH was slightly lower (0.62 *±* 0.09) compared to baseline LOR, but differences were not statistically significant. A summary of model performance can be found in Table 3. Confusion matrices for the top-performing models are shown in Figure 3H.

**Table 3:** Average model performance for pH and Apgar prediction tasks, trained and evaluated on the last 30 minutes of CTG. The standard error is shown in parentheses. Model performance was compared to clinician performance of 0.27 *±* 0.18 sensitivity at 90% specificity [36].

| Inputs | AUROC |  | Sensitivity at 90% Specificity |  |
| --- | --- | --- | --- | --- |
|  | pH | Apgar | pH | Apgar |
| Feature-based | 0.61 (0.06) | 0.35 (0.10) | 0.24 (0.14) | 0.00 (0.00) |
| UC | 0.55 (0.09) | 0.62 (0.11) | 0.13 (0.09) | 0.00 (0.00) |
| FHR | 0.54 (0.09) | 0.39 (0.09) | 0.17 (0.09) | 0.00 (0.00) |
| FHR + metadata | 0.55 (0.08) | 0.38 (0.16) | 0.06 (0.06) | 0.26 (0.20) |
| FHR + UC | 0.62 (0.09) | **0.69 (0.12)** | 0.27 (0.11) | **0.32 (0.20)** |
| FHR + UC + metadata | **0.69 (0.08)** | 0.57 (0.11) | **0.44 (0.13)** | 0.05 (0.12) |

**Figure 3.**
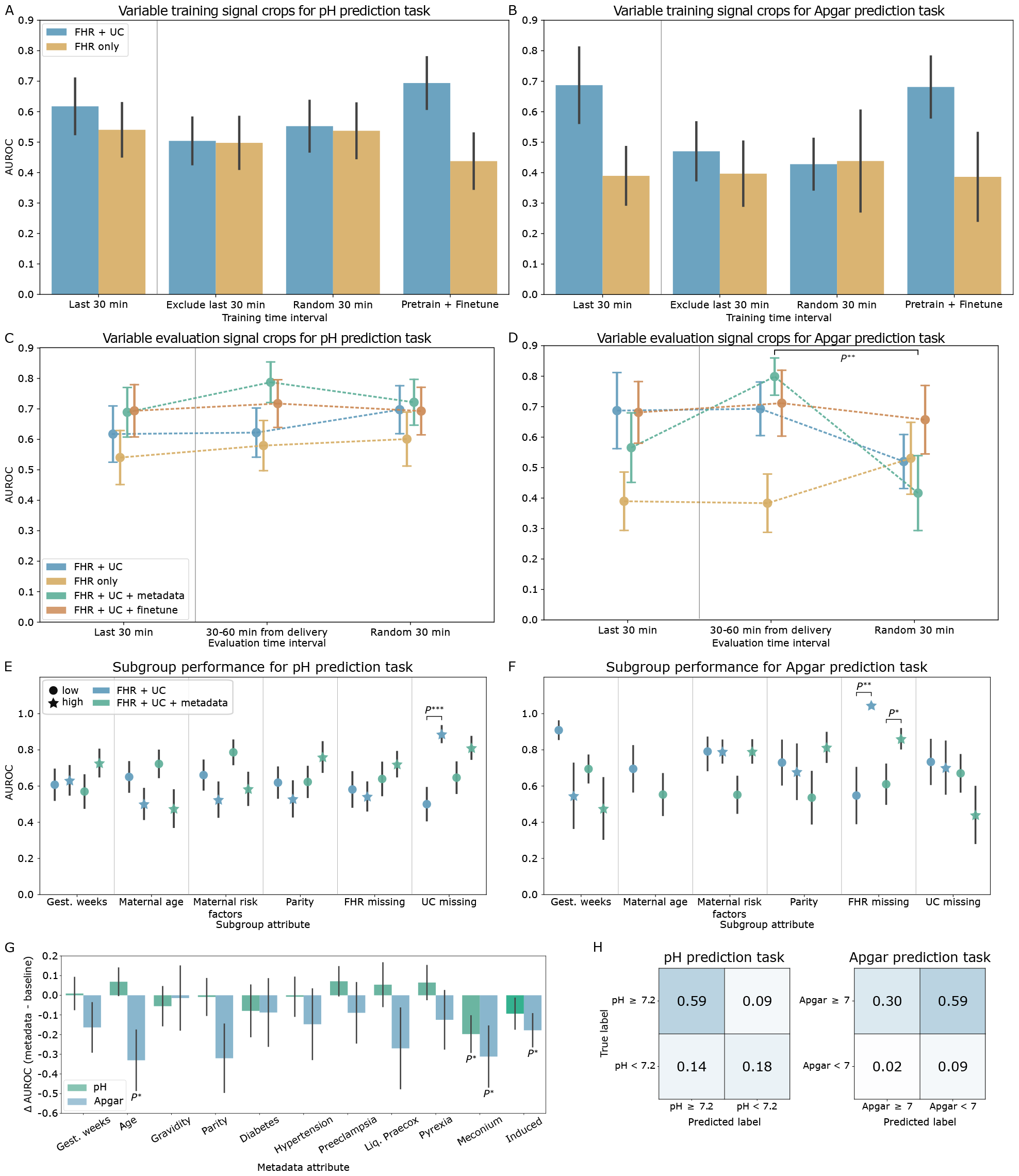
*P*^*∗*^ and *P*^*∗∗*^ represent a *p*-value *<* 0.05 and 0.01 respectively. Error bars depict the standard error. (A-D) AUROC for models trained on (A-B) and evaluated on (C-D) CTG signal from different time intervals of recording. Markers to the left of the vertical gray line indicate the paradigm used to train and evaluate the baseline models. (E-F) Subgroup AUROC performance for the FHR + UC and FHR + UC + metadata baseline models. Results for the high maternal age subgroup are omitted and further details can be found in Appendix C. Change in AUROC performance when adding a single metadata attribute to the baseline FHR + UC model input, compared to baseline. (H) Confusion matrix for the best-performing pH (*left*) and Apgar (*right*) prediction tasks.

#### Comparing deep learning and feature-based approaches for baseline model (FHR + UC)

For pH classification, the feature-based approach had a slightly lower, but non-significantly different, performance (0.61 *±* 0.06) than the CNN baseline (0.62 *±* 0.06). Apgar prediction, however, had a significantly reduced performance for the feature-based approach (0.35 *±* 0.10) compared to the CNN baseline (0.69 *±* 0.12).

#### Comparing FHR only, UC only, and FHR + UC

The FHR + UC model achieved the highest AUROC performance for both pH and Apgar classification tasks, followed by UC only and then FHR only models. Excluding either of the channels also resulted in a significant reduction in sensitivity at 90% specificity for both tasks.

#### Performance with maternal and fetal metadata

Adding metadata to the FHR + UC model increased the performance for the pH prediction task by 0.07 points to 0.69 *±* 0.08, though the results were not significant. Adding metadata for the Apgar prediction task degraded the FHR + UC model performance (0.57 *±* 0.11).

#### Identifying metadata contributions to model performance

For the pH prediction task, adding an indication of meconium presence in the amniotic fluid significantly decreased model performance. The addition of maternal age, pre-eclampsia status, premature rupture of membranes (liq. praecox), and pyrexia each contributed positively to model performance, but improvements were not statistically significant. For the Apgar classification task, the addition of metadata generally decreased performance. We observed a significant decrease in AUROC compared to the baseline FHR + UC model when including the maternal age, meconium, and induced delivery attributes. Full results are shown in Figure 3G.

### 3.2 Evaluation of temporal distribution shifts during training and testing

#### Training on different time points and evaluating on the last 30 minutes of the held-out set is shown in Fig. 3A-B

We observed no significant differences in AUROC from training on different time points and testing on the last 30 minutes for both pH and Apgar prediction tasks. However, Apgar prediction performance had higher variability across the different trained models compared to pH prediction performance, which was more stable. Pre-training on windowed signals before the last 30 minutes, then fine-tuning on the last 30 minutes achieved the highest AUROC for predicting pH using FHR + UC (0.69 *±* 0.09) followed by the model trained on the last 30 minutes alone (0.62 *±* 0.09). This agrees with clinical practice that the CTG signal recorded closest to delivery corresponds more with the recorded pH measurement. For the Apgar prediction task, pre-training and fine-tuning (0.68 *±* 0.10) the model achieves a similar performance as training on the last 30 minutes alone (0.69 *±* 0.12).

#### Training on the last 30 minutes and evaluating on different time points of the held-out set is shown in Fig. 3C-D

We observed no significant differences in the performance of the models when tested on signals across different time points for the pH classification task, simulating the evaluation setting in low-resource environments. In general, pH performance remained stable across different time point evaluations. Apgar prediction performance typically had higher variability across different time points, demonstrating reduced robustness to temporal distribution shifts.

### 3.3 Subgroup evaluation

Figures 3E and 3F show the AUROC performance of the subgroup analysis for pH and Apgar prediction, respectively. We found significant differences in baseline performance between subgroups with low and high UC signal missingness with pH evaluation and for FHR missingness subgroups with Apgar prediction.

With metadata, the performance disparities observed with pH prediction were mitigated. However, including metadata increased the AUROC performance disparities for demographic and clinical-related subgroups on this task, although none of these differences were statistically significant. This indicates that conditioning on demographic and clinical subgroup information may yield less equitable performance with limited sample sizes.

## 4 Discussion

Our study establishes the viability of employing deep learning for predicting fetal hypoxia from cardiotocography (CTG) tracings and the need for rigorous evaluations by choice of label, time interval, and subgroup performances. The baseline model, incorporating fetal heart rate (FHR) and uterine contraction (UC) signals, achieved performance levels consistent with prior work and clinical practice [19, 36]. For the pH prediction task, our findings indicate that including both FHR and UC channels is imperative for accurately identifying fetal hypoxia cases. Comparative analyses with different ground truth labels revealed that utilizing objective pH measurements yields more consistent performance than subjective clinician-assigned Apgar scores. Our research highlights the limitations of relying on Apgar scores as ground truth labels and encourages future work directed at predicting quantitative objective measures, such as umbilical cord blood pH. This insight is particularly pertinent for machine learning practitioners aiming to train machine learning models in low-resource medical settings lacking objective umbilical cord pH measurements [24, 25].

Furthermore, training models on the last 30 minutes of signal, the time interval most closely correlated with the delivery outcome, yielded the best performance. Notably, pre-training the model on signal data excluding this critical interval, followed by fine-tuning on the last 30 minutes, further improved performance. The robustness of our pH classification model to out-of-distribution time points was demonstrated by consistent performance across randomly sampled intervals within 90 minutes of delivery, simulating the intermittent CTG monitoring setting standard in LMICs. Subgroup analyses revealed performance disparities across demographic, clinical, and signal quality subgroups for the baseline model. However, while incorporating fetal and maternal metadata attributes during training enhanced pH classification performance, it exacerbated performance disparities for demographic and clinical subgroups.

In contrast to previous approaches [8–13], we leveraged deep learning to enable end-to-end prediction of fetal hypoxia, taking into account temporal and contextual cues often overlooked by feature-based methods. The robustness of our model highlights the compatibility of our approach with intermittent CTG monitoring settings, particularly in low-resource environments where intermittent monitoring is standard practice.

This study had several limitations that constrain the generalizability of our findings. First, we used CTGs from 552 patients at a single hospital in Prague, Czech Republic. To enhance the robustness of our findings, future investigations should involve a larger and more diverse dataset sourced from maternity centers worldwide, encompassing varied clinical contexts, demographics, and outcomes. Secondly, the absence of automated CTG digitization infrastructure in many resource-limited settings necessitates the simulation of intermittent CTG use cases from facilities with digitized recordings [37, 38]. Additionally, our study did not include a comparison of algorithmic performance against clinicians viewing the same dataset, prompting future research to explore different human and algorithmic use combinations. Finally, further work is needed to understand how such prediction algorithms can be optimally integrated into clinical workflows to improve neonatal outcomes.

## 5 Conclusion

We develop neural networks and feature-based models to interpret CTGs and propose a framework to evaluate these models. Our major findings indicate that utilizing objective pH measurements, as opposed to clinician-defined Apgar scores, results in more consistent, robust performance under temporal distribution shifts. This is especially important when transferring models to settings that only have intermittent CTG measurements. The model and evaluation framework we propose can be applied more generally to paired time-series datasets especially where sample size is limited.

## Data Availability

We used an open source dataset found at: https://physionet.org/content/ctu-uhb-ctgdb/1.0.0/
All additional data produced in the present study are available upon reasonable request to the authors

## Acknowledgments

The authors thank Stephen Pfohl and Chirag Nagpal for valuable discussions and feedback. This work was funded by Google.

## A Additional dataset details

### A.1 Preprocessing steps

A step-by-step depiction of the preprocessing pipeline is shown in Figure 4.

**Figure 4.**
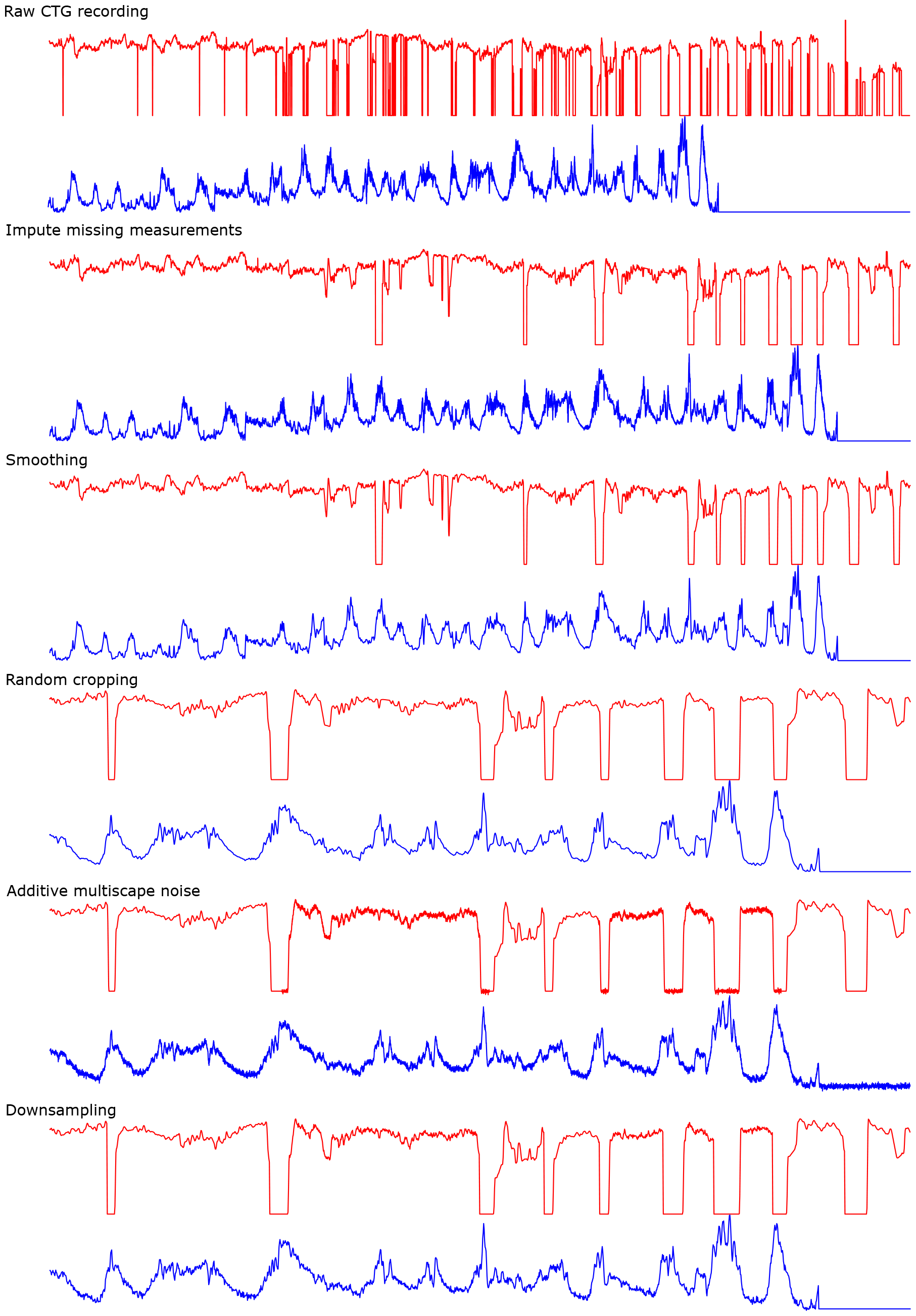
Preprocessing pipeline example.

#### Removal of repeated zero signals

Time points consisting of repeated zeros at the start and end of the FHR signal were removed from both the FHR and UC signals.

#### Data quality assessment

The remaining missing FHR measurements were flagged. A 5-minute sliding window with a 1-minute stride was used to ensure that the FHR signal loss was less than 50% within the window as described in Asfaw et al. [17]. Time steps that violate this condition were flagged for zero-value assignment after the imputation and smoothing steps. Missing FHR segments 15 seconds or longer were also flagged using a 15-second sliding window with a 0.25-second stride [23].

#### Imputation of missing values

Missing segments in both FHR and UC signals shorter than 15 seconds were imputed using linear interpolation. In the early stages of development, we also experimented with univariate XGBoost forecasting to interpolate missing segments longer than 15 seconds, but this method does not take the relationship between the FHR and UC into account and leads to decreased performance. All segments of longer duration were imputed with zeros.

#### Smoothing

Following Ogasawara et al. [19], both the FHR and UC signals were smoothed using a bilateral moving average with a rolling window length of 15 time points and a Hamming window.

#### Data augmentation and signal cropping

The preprocessed records were augmented using small-scale random cropping and additive multi-scale noise introduced in Zhou et al. [23] as a form of oversampling to mitigate the effects of class imbalance in the CTU-UHB dataset. This method performs small-scale shifts in the window to generate random crops of the last 30 minutes of the preprocessed record and adds either global or local Gaussian noise to the resulting cropped signal.

Ten randomly cropped records were generated for each preprocessed record by first extracting the starting time point for the last 30 minutes of signal and randomly sampling a new starting point up to 4 minutes before the original starting point. For each cropped record, we randomly sampled two binary variables from independent uniform Bernoulli distributions to denote whether to add local or global noise to each of the FHR and UC signals. If global noise was chosen, Gaussian noise with zero mean and a variance of 4 was added to the entire signal. If local noise was specified, we then sampled a number between 1 and 5 uniformly at random to specify the number of segments to add local noise. Finally, random start and end points for each of these segments were randomly sampled independently for each channel.

Although data augmentation was performed for all records, the augmented samples were only used to train the models. An evaluation dataset was constructed by deterministically cropping the record to the last 30 minutes of signal, skipping the random cropping and additive noise steps.

#### Downsampling

The resulting augmented and evaluation datasets were down-sampled from 4 Hz to 1 Hz to reduce the length of the input signal and remove redundant information since the average fetal heart beats less than 3 times a second (*<* 180 bpm).

### A.2 Rules-based feature extraction

Preprocessing steps up to and including the imputation of missing values described in the previous section were applied to generate a separate dataset to use with the feature extraction algorithm. Following the imputation of segments shorter than 15 seconds, the FHR and UC signals were smoothed with a rolling window of 120 time points (30 seconds), and missing segments longer than 15 seconds were imputed using linear interpolation. The same small-scale edge clipping and additive noise data augmentation process was applied to the smoothed signal.

The uterine contractions were identified by finding peaks in the UC signal that mark a bell-shaped gradual increase in the UC signal between 45 and 120 seconds in total duration. Adhering to the current International Federation of Gynecology and Obstetrics (FIGO) guidelines, we computed the FHR baseline over a 10-minute window [35]. The initial FHR baseline was the average value of the FHR signal within the window and the baseline was iteratively updated as accelerations and decelerations were identified and removed from the baseline calculation. This process was repeated until the magnitude of the FHR baseline change was less than 0.5. The baseline variability was computed by identifying the average amplitude change between successive small-scale peaks and troughs that represent baseline fluctuations. We identified FHR accelerations as increases in the FHR signal above baseline greater than 15 beats-per-minute (bpm) in amplitude, lasting more than 15 seconds, with the time from onset to peak shorter than 30 seconds and total duration less than 10 minutes.

Decelerations were identified as decreases in the FHR signal below baseline greater than 15 bpm in amplitude and lasting more than 15 seconds. We further categorized the decelerations according to the relation to the closest identified uterine contraction to the deceleration onset. Late decelerations are decelerations where the onset is more than 20 seconds after the corresponding UC onset but before the end of the UC. Decelerations not classified as late decelerations are denoted as early decelerations if the duration is shorter than 3 minutes, prolonged decelerations if between 3 and 5 minutes, and severe if longer than 5 minutes. Variable decelerations are those where the minimum value of the deceleration is less than 30 seconds from the deceleration onset.

### A.3 Alternative time interval datasets

We generate pre-training samples by systematically cropping the available preprocessed signal excluding the last 30 minutes of the signal using a 30-minute sliding window with a 1-minute stride. The number of samples generated for each record scales with the duration of the preprocessed record, so an unequal number of samples are generated for different record identifiers. Additive multiscale Gaussian noise was applied to each of these cropped samples to generate 5 augmented samples for each cropped sample.

The randomly cropped dataset consists of 30-minute segments cropped over the entire signal. This dataset is partitioned into an augmented dataset, with one noisy sample per cropped sample, and an evaluation dataset with no data augmentation.

Lastly, we consider the evaluation dataset from the 30 minutes of the signal immediately before the last 30 minutes of the signal. Record identifiers with a preprocessed signal length shorter than 60 minutes were excluded from this dataset due to a lack of available signal.

### A.4 Training, validation, and test sample sizes

The number of preprocessed CTG recordings in each training, validation, and test split for the various datasets used to train and evaluate the neural network models is summarized in Table 4. All training datasets had an equal distribution over the binary class label. For the dataset with the last 30 minutes of signal, the proportion of abnormal samples was 0.32 and 0.32, 0.12 and 0.11, and 0.40 and 0.32 for the validation and test sets for the pH, Apgar, and LOR prediction tasks respectively.

**Table 4:**
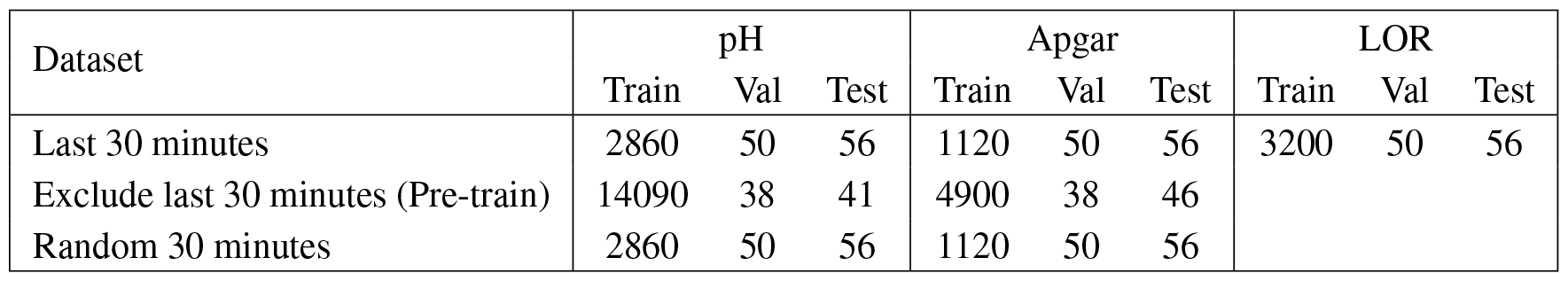
Number of preprocessed CTG recordings for each dataset and prediction task combination.

| Dataset | pH |  |  | Apgar |  |  | LOR |  |  |
| --- | --- | --- | --- | --- | --- | --- | --- | --- | --- |
|  | Train | Val | Test | Train | Val | Test | Train | Val | Test |
| Last 30 minutes | 2860 | 50 | 56 | 1120 | 50 | 56 | 3200 | 50 | 56 |
| Exclude last 30 minutes (Pre-train) | 14090 | 38 | 41 | 4900 | 38 | 46 |  |  |  |
| Random 30 minutes | 2860 | 50 | 56 | 1120 | 50 | 56 |  |  |  |

### A.5 Subgroup definition criteria

Subgroups were split on a binary subgroup variable and held-out CTG recordings were assigned to two disjoint sets according to a cut-off threshold value, shown in Table 5. The thresholds were chosen according to clinical understanding [39–41]. For the subgroup analysis, the low and high groups consisted of recordings below and above the threshold respectively.

**Table 5:**
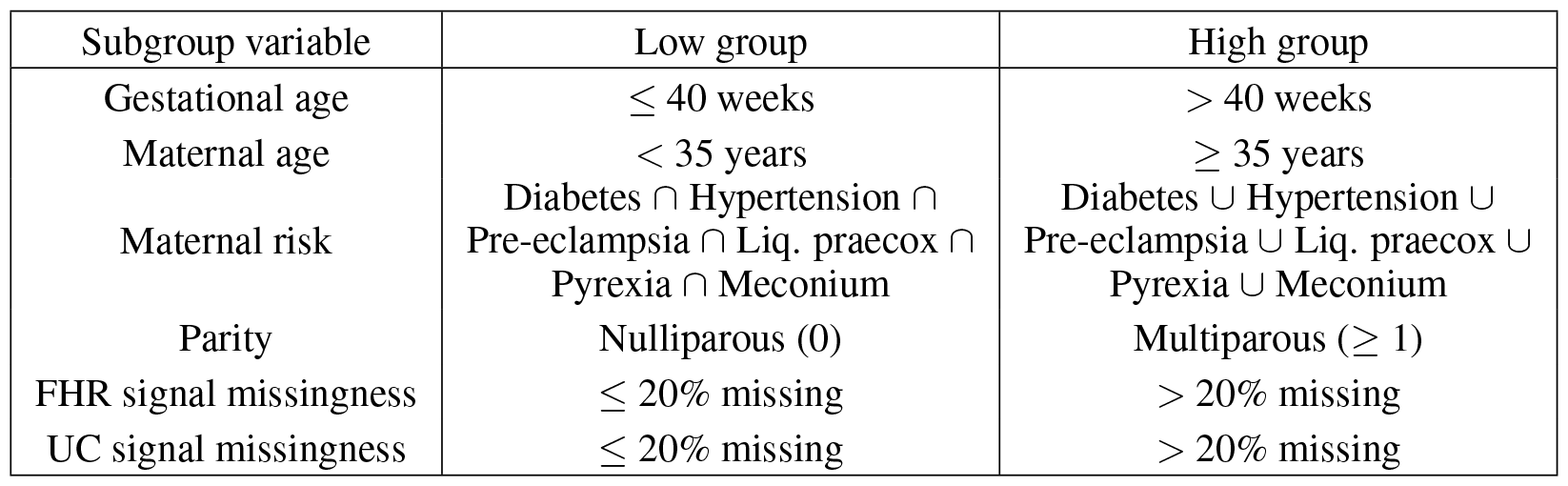
Binary demographic, clinical, and signal quality subgroup variable cut-off thresholds.

### A.6 Signal quality descriptors

Signal quality metrics were computed after the cropping step in the preprocessing pipeline and are summarized for each time interval in Table 6.

**Table 6:** Average proportion of missing signal in the FHR and UC channels and the number of records (out of 552) that contain high (*>* 20%) signal missingness for each evaluated dataset.

| Dataset | Mean |  | Count (out of 552) |  |
| --- | --- | --- | --- | --- |
|  | % FHR missing | % UC missing | High FHR missing | High UC missing |
| Last 30 minutes | 0.143 | 0.143 | 147 | 182 |
| 30-60 minutes from delivery | 0.116 | 0.117 | 84 | 86 |
| Random 30 minutes | 0.117 | 0.122 | 214 | 226 |

## B Hyperparameter tuning

To optimize the neural network model hyperparameters, we performed an architecture search over the number of temporal, depthwise, separable filters, the kernel width for the separable convolution, the number of hidden layers, and the hidden layer dimension with fixed model training hyperparameters (loss function: binary cross-entropy, learning rate: 3e*−*4, batch size: 128, dropout: 0.2). The range of values considered and the sampling function used to conduct the model architecture and training hyperparameter search is depicted in Table 7. The resulting optimized model architecture parameters and fixed hyperparameter values used to conduct the architecture search are shown in Table 8.

**Table 7:**
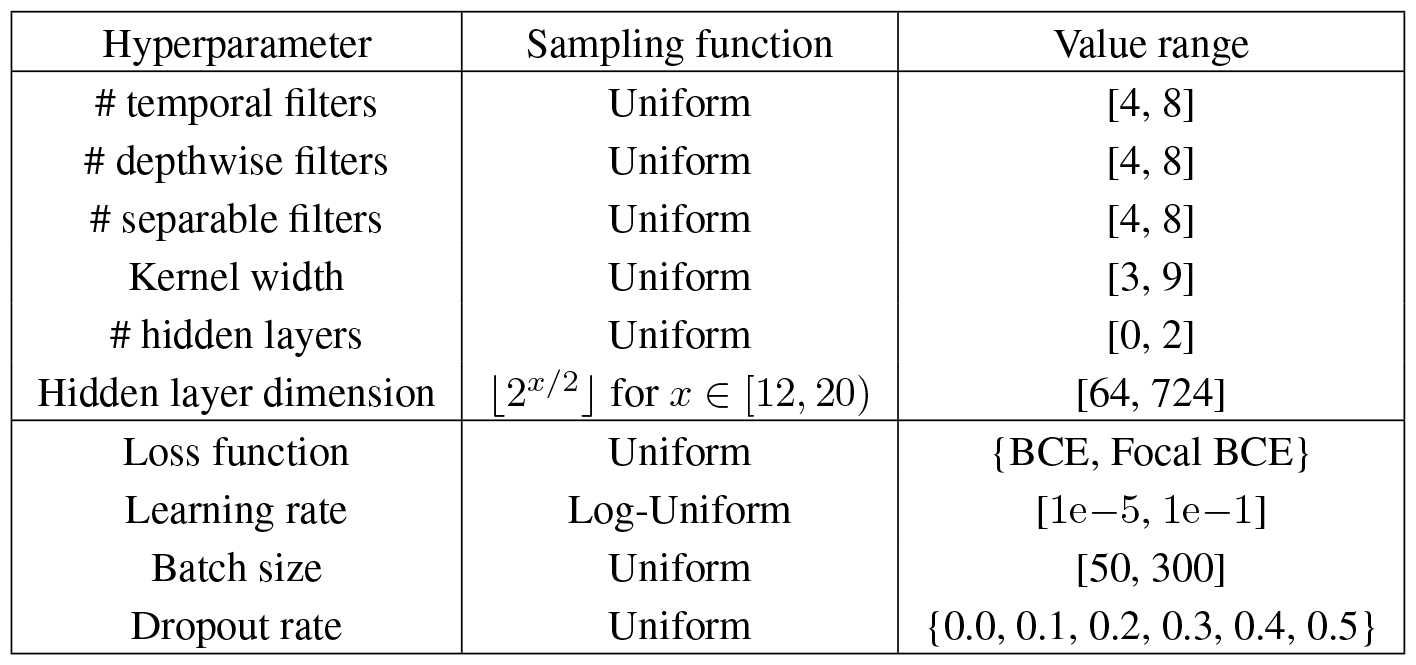
Model architecture and training hyperparameter search sampling parameters.

**Table 8:** *Top*: Selected model architecture hyperparameters. *Bottom*: Default model training hyperparameters used during the architecture search.

| Hyperparameter | Default value |  |
| --- | --- | --- |
|  | 2-channel | 1-channel |
| # temporal filters | 6 | 7 |
| # depthwise filters | 8 | 6 |
| # separable filters | 5 | 4 |
| Kernel width | 5 | 3 |
| # hidden layers | 2 | 2 |
| Hidden layer dimension | 128 | 181 |
| Loss function | BCE |  |
| Learning rate | $3e-4$ | |
| Batch size | 128 |  |
| Dropout rate | 0.2 |  |

We generated confidence intervals for the estimated metrics by performing 1000 iterations of bootstrap resampling of the 10% held-out test set, corresponding to a bootstrap size of 56. As mentioned in Section 2.4, we selected the best hyperparameter settings using the highest validation AUROC averaged over cross-validation folds. We then selected the best-trained model overall by choosing the model trained on the cross-validation fold yielding the highest validation AUROC. The same procedure was used to select the model from the pre-training phase to use for downstream fine-tuning.

## C Invalid performance metrics

The maternal age subgroup performance metrics for the Apgar prediction task were not comparable because only the normal class was present in the test set for the high maternal age subgroup. This yielded invalid AUROC and sensitivity metrics. For this subgroup, none of the 10 records in the held-out test set had an associated abnormal Apgar score (mode: 9, median: 9, min: 8). However, both the FHR + UC and FHR + UC + metadata models predicted all normal Apgar scores for this subgroup, achieving perfect accuracy. We speculate that the model defaults to predicting normal Apgar scores due to the class imbalance in the dataset. This may lead to an over-optimistic estimate of performance on the test set since the high maternal age subgroup has an under-representation of abnormal Apgar scores. Therefore, the model’s ability to identify abnormal cases for this subgroup remains yet to be evaluated.

